# Computational prediction of therapeutic response and cancer outcomes

**DOI:** 10.1101/2024.01.17.24301444

**Authors:** Matthew Griffiths, Amanzhol Kubeyev, Jordan Laurie, Andrea Giorni, Luiz A. Zillmann da Silva, Prabu Sivasubramaniam, Matthew Foster, Andrew V. Biankin, Uzma Asghar

## Abstract

Oncology therapeutic development continues to be plagued by high failure rates leading to substantial costs with only incremental improvements in overall benefit and survival. Advances in technology including the molecular characterisation of cancer and computational power provide the opportunity to better model therapeutic response and resistance. Here we use a novel approach which utilises Bayesian statistical principles used by astrophysicists to measure the mass of dark matter to predict therapeutic response. We construct “Digital Twins” of individual cancer patients and predict response for cancer treatments. We validate the approach by predicting the results of clinical trials. Better prediction of therapeutic response would improve current clinical decision-making and oncology therapeutic development.

## Introduction

Therapeutic development in oncology continues to be challenging. Whilst significant advances have been made in some instances, progress continues to be slow and incremental. The vast majority of candidate therapies fail, and the failure rate in advanced phase 3 clinical trials remains high. This inefficiency costs over $50 billion per annum, which is unsustainable for most health systems and economies.

Advances in the molecular profiling of cancer, coupled with accelerated computing power, provide the promise of moving away from a “trial and error” approach to cancer treatment and therapeutic development, to one where we can predict therapeutic efficacy prior to treatment. “Digital Twins”; in-silico virtual replicas of cancer patients, offer enticing possibilities for improving cancer treatment. The benefits of accurate prediction of therapeutic response and patient outcome can be applied at many points in therapeutic development, from early candidate drug selection through to late phase clinical trials and routine cancer care.

Here, we present a machine learning approach which simulates treatment with cytotoxic and small molecule therapies with applicability across numerous cancer types. We demonstrate how these models can predict overall response rates (ORR) for a range of cancer types and treatments. The digital twins predict drug efficacy, for single agent or drug combinations and can predict if treatment A will perform better than treatment B in individual patients and in virtual clinical trials. The prediction accuracies were tested against actual response rates and overall survival metrics in historical clinical trials. Synthetic controls for comparator arms of clinical trials were constructed to enable benchmarking of predicted clinical efficacy of investigational drugs versus standard of care. The simulated clinical trial can then predict survival. Finally, we demonstrate how this approach can be used for patient cohort enrichment for an investigational drug of interest, and calculate the predicted increase in response rates achieved through such enrichment strategies.

## Results

### Constructing Digital Twins to simulate therapeutic response and clinical trials

The modelling approach we used arose out of a collaboration with astrophysicists^1^ to develop advanced Bayesian inference software that enables integrative modelling of gravitational lensing and cancer biology. These partnerships motivated a transfer learning approach where detailed molecular and therapeutic data generated from biological experimentation was used to build generalisable Bayesian models that can be applied to predict treatment efficacy for single agents and combinations.

We created a computational framework that could predict in vitro therapeutic response and clinical response and survival using multi-dimensional data that included molecular profiles, predominantly genomic and transcriptomic. Digital twins were created to address specific questions using 3 distinct models: 1) Drug Efficacy Model 2) Treatment Response Model and 3) Overall Survival Model (Figure 1).

**Figure 1:**
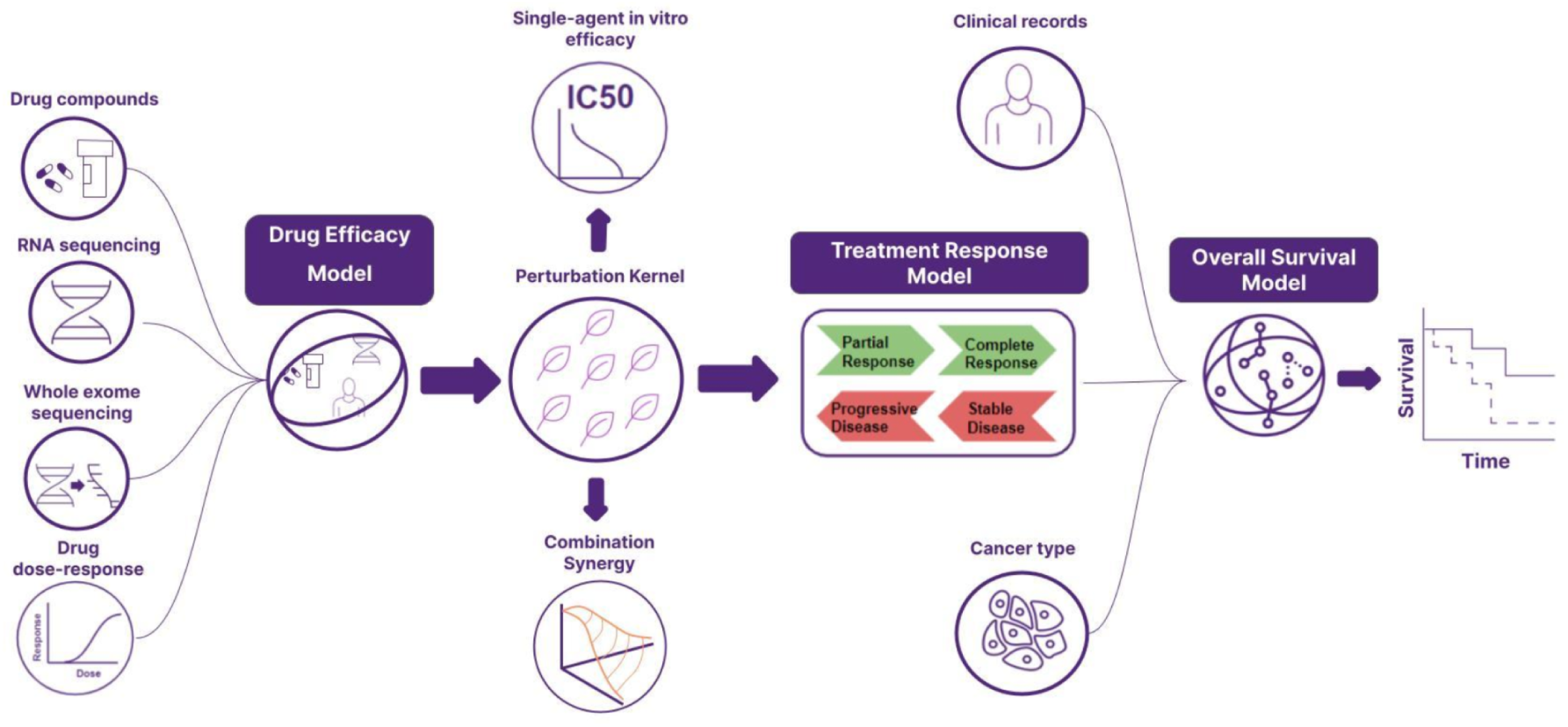
Schematic of the Digital Twin Simulator designed to model in silico therapeutic response and clinical trials. The components that underpin the digital twin are the Drug efficacy, the Treatment response and the Overall survival models. **The Drug Efficacy Model** can ingest pre-clinical and/or clinical data. It uses the molecular profiles of tumours or preclinical models such as gene expression and mutation profiles and a drug’s molecular fingerprint derived from the compound structure. The drug efficacy model generates a perturbation kernel, which calculates the similarity of the effect of a drug perturbation between two samples treated with two drugs. Gaussian process regression using this kernel can predict multiple types of treatment response predictions such as *in vitro* IC_50_ and drug synergy scores, and provides inputs for the Treatment response model. The **Treatment Response Model** predicts patient response with outputs as two states: either response (partial response or complete response using RECIST) or no response (stable disease or progressive disease). The Treatment response model provides input for the Overall Survival model. The **Overall Survival Model** integrates inputs from individual patient clinical data, treatment response, pathology and if available molecular profiles (gene expression +/- mutation profiles +/- copy number alterations). The Overall Survival Model predicts overall survival for individual patients given a specific treatment regimen and can be modified to consider alternative endpoints such as disease-free survival (DFS) to reflect clinical trial study endpoints.

The perturbation kernel is derived from the drug-efficacy model and is leveraged by multiple Bayesian inference models to transfer understanding about shared molecular mechanisms across *in vitro* combination screens and clinical treatment settings. It defines the similarity in the molecular mechanisms between every pair of datapoints (for example a patient treated with a taxane such as docetaxel vs. a different patient treated with an anthracycline such as doxorubicin), which can be used to make predictions of effect using Gaussian processes^2^.

The drug efficacy and perturbation kernel were built using *in-vitro* dose-response data from the Cancer Therapeutic Response Portal (CTRP)^3–5^. The CTRP dataset consists of 481 anti-cancer compounds which include chemotherapy and targeted small molecules. These were dosed against 860 cancer cell lines. The molecular data for the cell lines was obtained from the Cancer Cell Line Encyclopedia^6^. This dataset was used to train the perturbation kernel to predict IC_50_ for the compounds in the dataset using a Sparse Gaussian Process. The perturbation kernel can also accurately predict synergy scores from the NCI-ALAMANAC dataset (unpublished data) for combination treatments. In this study, the model was tested using the perturbation kernel to predict treatment response in clinical data using the TCGA dataset located at the NCI Genomic Data Commons^7^. A summary of the datasets used and abbreviations can be found in Table 1 and Table 2. A detailed breakdown of the cohorts used in this study can be found in Extended Data Figure 1.

**Table 1.**
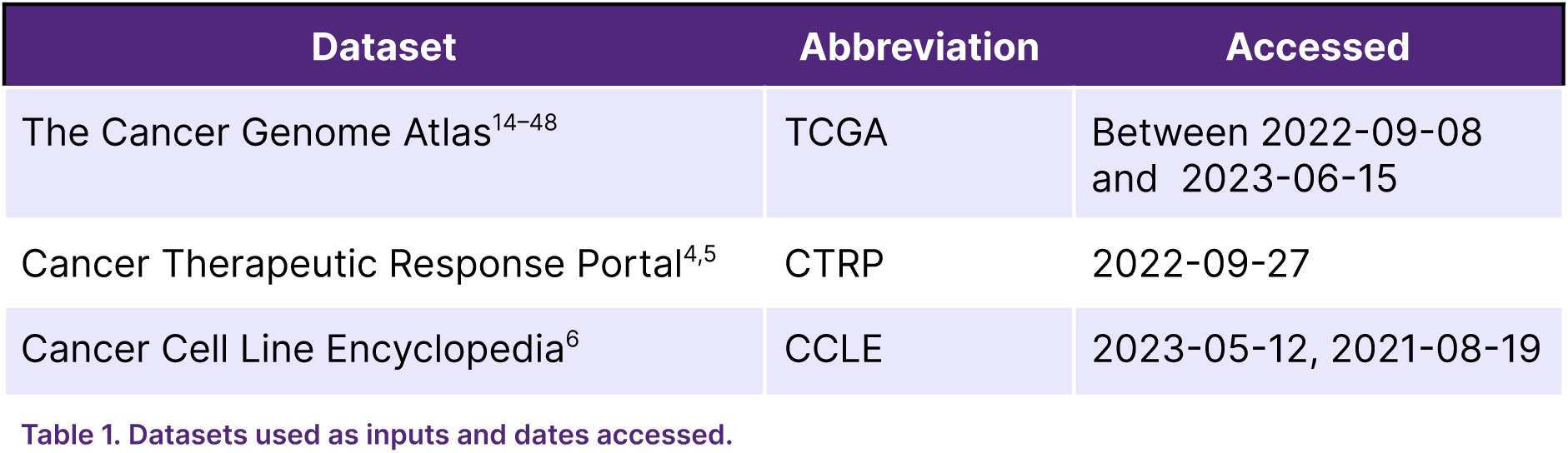
Datasets used as inputs and dates accessed.

| Dataset | Abbreviation | Accessed |
| --- | --- | --- |
| The Cancer Genome Atlas<sup>14-48</sup> | TCGA | Between 2022-09-08 and 2023-06-15 |
| Cancer Therapeutic Response Portal<sup>4,5</sup> | CTRP | 2022-09-27 |
| Cancer Cell Line Encyclopedia<sup>6</sup> | CCLE | 2023-05-12, 2021-08-19 |

**Table 2:**
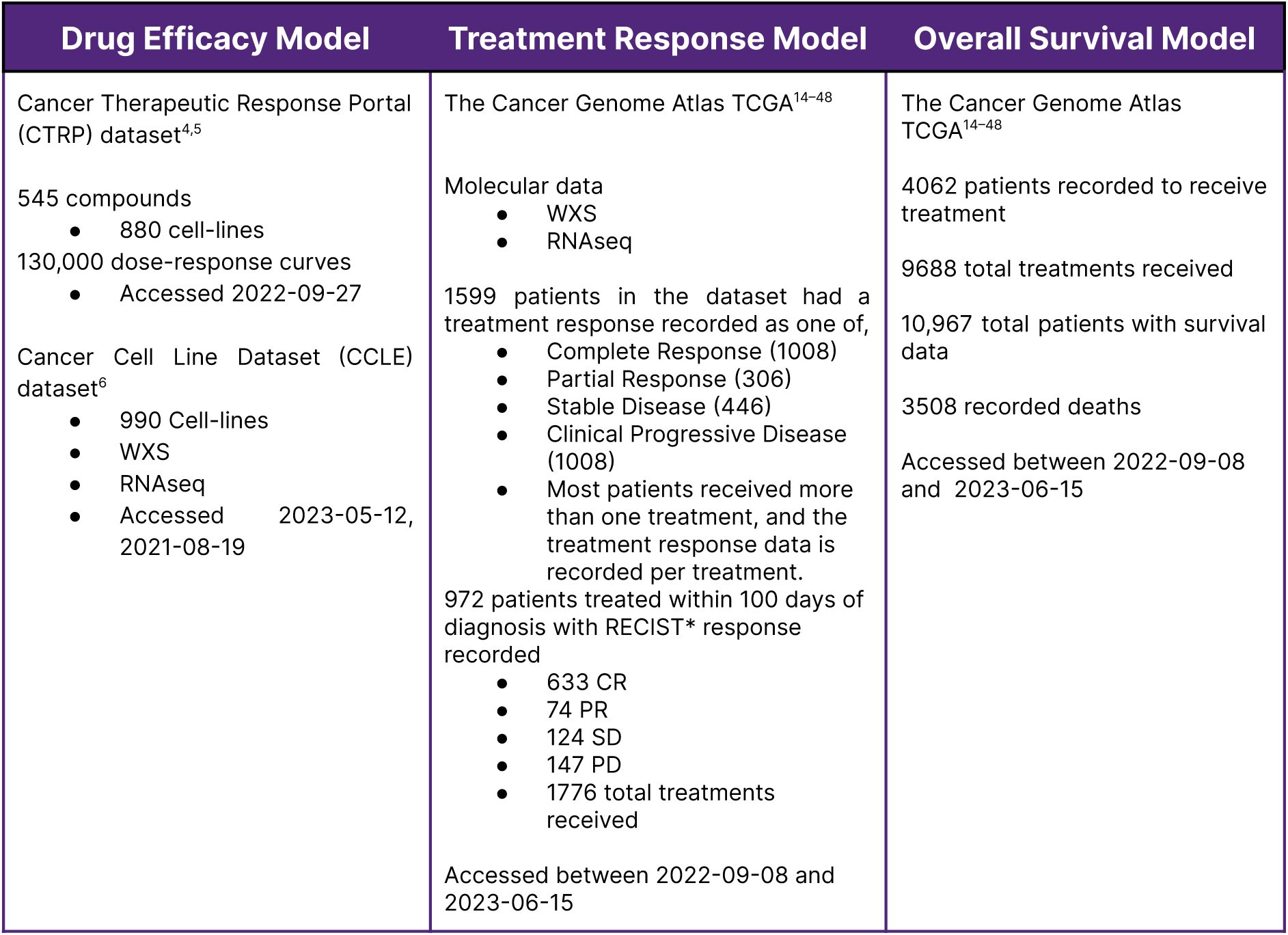
Input data for predictive modelling. WXS, whole exome DNA sequencing; CR, complete response; PR, partial response; SD, stable disease; PD, progressive disease; RECIST - Response Evaluation Criteria in Solid Tumours^49^.

To evaluate the performance of the model across the TCGA dataset we split the dataset into 5 cross-fold splits, stratified by cancer type and overall survival. We then trained the models on 4 of the splits and predicted outcomes for the remaining (omitted) split. All accuracy metrics reported are averages of the metrics calculated for each split in turn. Many patients had missing information, these missing variables were either imputed by the mean value of that column for that patient’s cancer type or from the entire cohort. These mean values were calculated only from the training cross folds when imputing for the validation cohort. The treatment response model was used to calculate treatment response probabilities for all the patients using the training cross-folds. If the patient received no treatment, then the patient was considered to have 0 probability of treatment response.

The model used molecular fingerprints generated by the CDK^8–11^ and Cinfony^12^ chemoinformatics libraries from canonical SMILES structures obtained from PubCHEM^13^ to incorporate structural information about each therapeutic. This process restricts the treatment response predictions to small molecule therapies at this time and hence we focus on chemotherapy drugs as monotherapy and drug combinations.

### Validation of Digital Twin predictions using clinical trial simulations

We simulated digital trial arms for single chemotherapy drugs and combinations to predict treatment response in cancer patients with the goal of assessing the accuracy of digital twin predictions through comparison to historical clinical trial results. TCGA data was used as input, no original individual participant clinical trial data is available and was not used for predictions. Initially, the digital twin predictions were evaluated using an unblinded approach where our technology team was aware of the results. (Figure 2). The model treatment predictions were compared to the results of four historical phase 2 and phase 3 clinical studies (1997 - 2018). These were trials in metastatic pancreatic cancer (Burris, et al.^50^), advanced breast cancer (Chan, et al.^51^ and Tutt, et al.^52^) and platinum-sensitive recurrent ovarian cancer (Cantù, et al.^53^). We compared the predicted log odds ratio (OR; Figure 2a) generated by the digital twin model for Overall Response Rate (ORR) for each treatment arm tested in the clinical study, and then compared this against the actual reported log odds ratios (log OR) from the historical trial, the ground truth (Figure 2b). We started with single-agent predictions, then progressively increased the complexity through combinations and heterogenous treatments in more sophisticated clinical trial designs (Table 3).

**Figure 2:**
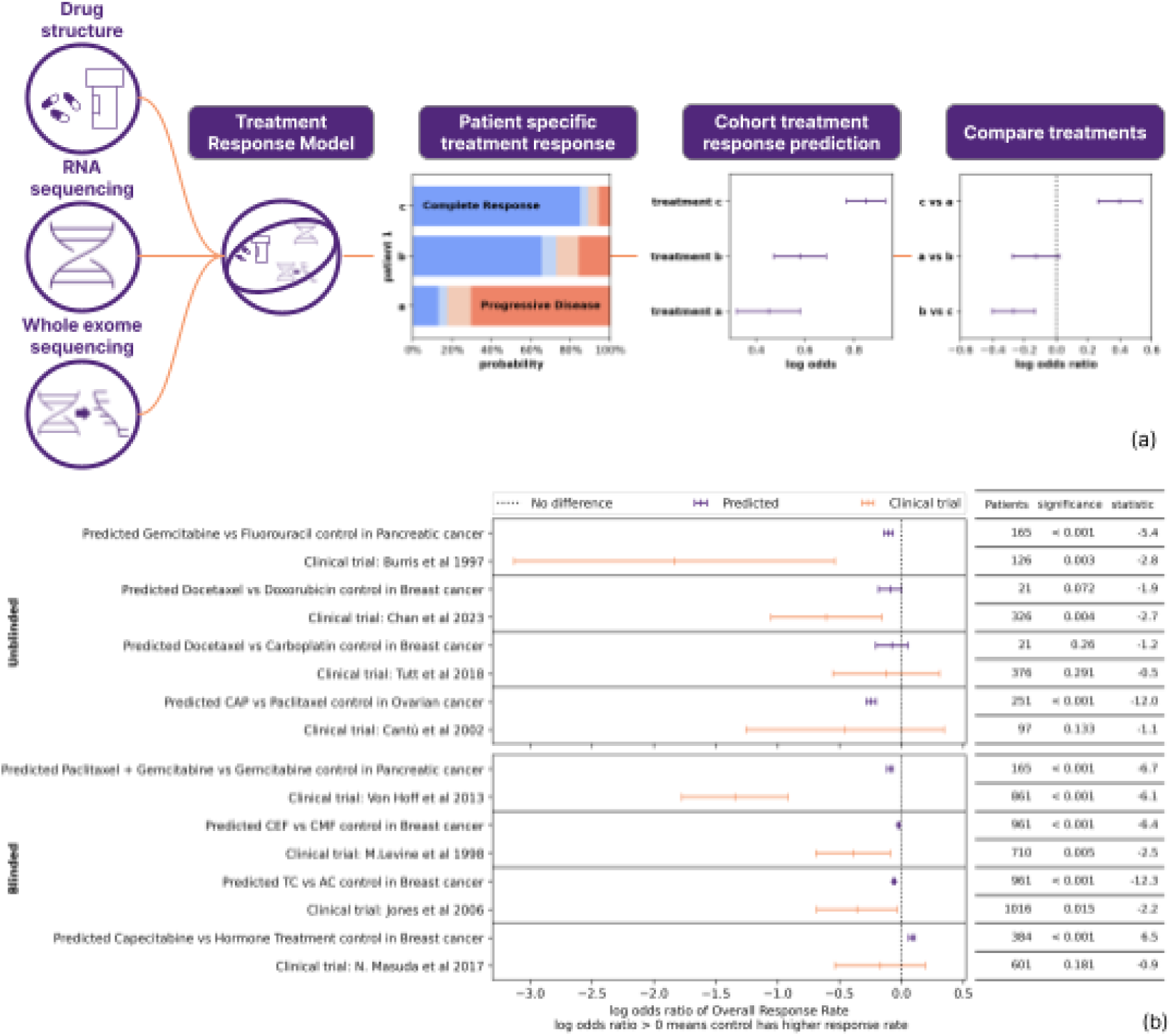
Clinical Trials simulations by Digital Twin model (Unblinded and blinded). The predictions from simulations of eight clinical trials by Digital Twins are shown by comparing the control arm and investigational arm, and predicting the difference in drug efficacy. Model accuracy was tested by a comparison of the predicted log odds ratio log(OR) for Overall Response Rate (ORR) by the model against the actual log Odds Ratios reported from clinical trials^50–53^. For metastatic/advanced cancer studies, the drug response rate was calculated using complete response + partial response. For adjuvant cancer studies, in the absence of in situ primary cancer, drug response was calculated by defining clinical response as the absence of disease relapse at a specified time point, and lack of response is equivalent to cancer relapse. 95% confidence intervals are shown for each log(OR) value; Digital Twin predictions (purple) and actual reported Clinical trial outcomes (orange). The threshold is set at zero, where >0 suggests the control arm has a better response and < 0 suggests that the investigational arm is better. The number of patients used to generate predictions or recruited into the study is reported on the right, with significance and statistics, see <u>Statistics</u> section in the Online Methods sections for details on how these values were calculated. The numbers used to make these plots are shown in Table 3.

**Table 3.** Summary of clinical trials compared to predictions. Abbreviations: ORR=Overall Response Rate; OS=Overall survival; mOS= median Overall Survival; DFS=Disease free survival; CAP= Cyclophosphamide, Doxorubicin and Cisplatin, ER+=Estrogen receptor positive, HER2-ve=human epidermal growth factor receptor 2 negative

| Comparison |  |  |  |  |  |  | Predicted |  |  |  |  |  |
| --- | --- | --- | --- | --- | --- | --- | --- | --- | --- | --- | --- | --- |
| Study | Trial | Patients | log Odds Ratio | log Odds Ratio SE | significance | statistic | Patients | log Odds Ratio | log Odds Ratio SE | significance | statistic | Proportion better outcome |
| Unblinded Validation |  |  |  |  |  |  |  |  |  |  |  |  |
| Burris et al 1997 | Metastatic Pancreatic Cancer Median survival 5.65 vs. 4.41 months for Gemcitabine compared to 5-FU, (P = .0025) | 126 | -1.833 | 0.661 | 0.003 | -2.77 | 165 | -0.105 | 0.019 | 2.64E-07 | -5.371 | 65.5% |
| Chan et al 2023 | Metastatic breast cancer. ORR Docetaxel 47.8% vs. Doxorubicin 33.3% P =.008) | 326 | -0.606 | 0.228 | 0.004 | -2.65 | 21 | -0.090 | 0.048 | 7.25E-02 | -1.896 | 66.7% |
| Tutt et al 2018 | First line treatment in metastatic Triple Negative breast cancer ORR 31.4% for Carboplatin vs. 34.0% for Docetaxel; P = 0.66) | 376 | -0.121 | 0.220 | 0.291 | -0.55 | 21 | -0.076 | 0.066 | 2.60E-01 | -1.159 | 71.4% |
| Cantù et al 2002 | Recurrent ovarian cancer. ORR 55% vs. 44.7% for CAP vs. Paclitaxel. (P = 0.062) | 97 | -0.455 | 0.409 | 0.133 | -1.11 | 251 | -0.243 | 0.020 | 1.38E-26 | -12.025 | 76.9% |
| Blinded Validation |  |  |  |  |  |  |  |  |  |  |  |  |
| Von Hoff et al 2013 | Metastatic Pancreatic Cancer. OS 8.5 months for gemcitabine/nab-paclitaxel group vs. 6.7 months for gemcitabine alone (HR 0.72, p<0.001) | 861 | -1.345 | 0.219 | 0.000 | -6.15 | 165 | -0.090 | 0.013 | 2.67E-10 | -6.732 | 32.1% |
| M.Levine et al 1998 | Adjuvant breast cancer. Disease-free survival for CMF (control arm) 53% vs 63% for CEF (p=0.009) | 710 | -0.389 | 0.153 | 0.005 | -2.55 | 961 | -0.023 | 0.003 | 1.85E-10 | -6.443 | 26.7% |
| Jones et al 2006 | Breast cancer. DFS for TC 86% vs. 80% for AC (HR 0.67; P = 0.015) | 1016 | -0.359 | 0.166 | 0.015 | -2.16 | 961 | -0.059 | 0.005 | 2.76E-32 | -12.277 | 45.6% |
| N. Masuda et al 2017 | Adjuvant breast cancer, addition of capecitabine to standard of care. ER+ve HER2-ve subgroup HR 0.73 for OS (95% CI 0.38-1.40; P=0.41) | 601<br>ER+ve<br>/910 | -0.170 | 0.186 | 0.181 | -0.91 | 384 | 0.087 | 0.013 | 2.66E-10 | 6.490 | 38.3% |

### Unblinded validation

#### Single-agent chemotherapy arms

##### Study 1

The first clinical trial we simulated reported by Burris et al. in 1997, was a prospective, randomised clinical trial in advanced pancreatic cancer^50^ (n=126; 17 sites in USA & Canada; 1997), which randomised participants to either single-agent 5-fluorouracil (n=63) or gemcitabine (n=63). Clinical benefit was 23.8% for gemcitabine compared with 4.8% for 5-fluorouracil (5-FU) (P = 0.002). Median survival was 5.65 months for gemcitabine compared to 4.41 months for 5-FU (P = .0025). Survival at 12 months was 18% for gemcitabine and 2% for 5-FU. Both chemotherapy drugs are considered to be anti-metabolites, therefore this experiment tested the model’s ability to detect the difference in drug efficacy for two drugs belonging to the same drug class. The Digital Twin drug response predictions were based upon either no response (stable disease or disease progression) or response (partial response or complete response). The model correctly predicted gemcitabine chemotherapy would have greater clinical benefit than 5-FU (predicted log odds ratio −0.10, P < 0.0001)(Figure 2).

##### Study 2

Two metastatic breast cancer studies were simulated, both designed to prospectively compare single-agent therapeutic arms. These studies considered either an anthracycline, a taxane or a platinum. Chan et al.^51^ reported a prospectively randomised phase 3 study comparing taxane monotherapy (docetaxel; n=161) vs. anthracycline monotherapy (doxorubicin; n=164) in metastatic breast cancer (n=326) previously treated with an anthracycline-containing regimen (UK & Europe). The Digital Twin model predicted single-agent docetaxel would be a better treatment than single-agent doxorubicin using a relatively small dataset of n=21 (predicted log odds ratio −0.09, P = 0.07)(Figure 2 and Table 3) The borderline P value reflecting the small number of patients available for prediction expanding the confidence interval. The actual trial (n=326) showed that docetaxel had a higher objective response rate than doxorubicin (47.8% vs. 33.3%; P =.008).

##### Study 3

The other Phase 3 Clinical study in breast cancer reported by Tutt et al.^52^ (TNT; 17 sites, UK) compared carboplatin (n=188) vs. a taxane, docetaxel (n=188) as first-line treatment in metastatic triple negative breast cancer (n=376). This study showed that carboplatin was no more active than docetaxel in the BRCA wildtype subpopulation of patients (ORR, 31.4% vs. 34.0%, respectively; P = 0.66). The Digital twin model predicted, in alignment with the trial results, that neither carboplatin nor docetaxel would have superior efficacy in the BRCA wild-type population. In summary, the Digital twin model accurately predicts chemotherapy responses for different drug classes and can effectively predict the difference in drug activity, if present for single-agent treatments.

#### Single-agent *vs.* combination chemotherapy in relapsed disease

##### Study 4

To continue to ascertain the potential limitations of the Digital Twin model, we then added complexity to the predictions we set by including combination treatments and used a study comparing a single drug against a drug combination. The Digital Twin model virtually simulated a prospective randomised study in ovarian cancer reported by Cantù et al. 2022^53^ (n=97; Italy) allocating participants to either single agent paclitaxel (taxane; n=50) or a combination of cyclophosphamide (alkylating agent), doxorubicin (anthracycline) and cisplatin (platinum) (CAP; n=47). This study recruited patients with recurrent ovarian cancer who had achieved complete remission with previous platinum-based regimens, and whose disease recurred after a progression-free interval of more than 12 months. The Digital Twin model used data inputs from the TCGA cohort to predict that the cisplatin-based combination (CAP) would have a higher response rate than paclitaxel. The Digital twin prediction reflected the actual published higher overall treatment response rate for CAP, which was 55% vs. 44.7% for CAP vs. Paclitaxel. (P = 0.062). Predicted ORR log odds ratio −0.24, P < 0.001 for n= 251 vs. calculated log odds ratio from clinical trial data −.044, P=0.133).

#### Blinded validation

As part of the validation process, the technology team were blinded to the published outcomes of an additional four phase 3 clinical trials. Three clinical studies in early breast cancer evaluated adjuvant chemotherapy regimens^54–56^ and one evaluated first-line metastatic pancreatic cancer^57^ (Figure 2). The Digital Twin model correctly predicted drug efficacy and the clinical trial result for all four clinical studies.

##### Study 5 (USA, Europe & Australia)

Von Hoff et al.^57^ The phase III Metastatic Pancreatic Adenocarcinoma Clinical Trial (MPACT) in metastatic pancreatic cancer compared the combination of nab-paclitaxel plus gemcitabine vs. gemcitabine alone as first-line therapy. The study randomised 861 previously untreated metastatic pancreatic cancer patients between these treatment arms. The response rates were 23% for nab-paclitaxel plus gemcitabine versus 7% for gemcitabine alone (P<0.001). Median overall survival was 8.5 months in the nab-paclitaxel-gemcitabine group vs. 6.7 months with gemcitabine alone (hazard ratio 0.72, p<0.001). Using data for a similar chemotherapy drug, paclitaxel, the Digital Twin model was able to correctly predict that nab-paclitaxel plus gemcitabine was superior to gemcitabine alone (Predicted log odds ratio −0.090, p = <0.001) (Figure 2).

##### Study 6 (National Cancer Institute of Canada Clinical Trials Group)

In order to test the model’s limitations with regard to the number of patients the model needed to train on, we designed a virtual trial with methotrexate chemotherapy, because the model had trained on only eight cancer patients treated with methotrexate. The study was a Phase 3 prospective randomised trial reported by Levine et al.^55^ (1998), which enrolled high-risk, node positive pre/peri-menopausal women post mastectomy or lumpectomy and axillary dissection (n=716) and randomised them to either adjuvant ECF (epirubicin, cyclophosphamide and fluorouracil), or adjuvant CMF (cyclophosphamide, methotrexate and fluorouracil) treatment. The relapse-free survival for CMF (control arm) was 53% (95% CI, 45-58%) and 63% (95% CI, 57-68; P=0.009) for CEF at 5 years. Although CEF was a more effective chemotherapy regimen, it was associated with significantly more acute toxicities and as a consequence is not widely used. In order to predict treatment response in the adjuvant setting where the cancer has been surgically removed. Virtual simulations by the digital twin model accurately predicted that adjuvant ECF would be superior to adjuvant CMF in early breast cancer (Predicted log odds ratio −0.023, P =<0.001; Table 3).

##### Study 7 (USA)

Reported by Jones et al. 2006^56^ with 1016 participants with a median follow up of 5.5 years, a phase 3 prospective randomised trial in stage 1-3 breast cancer reported disease-free survival at 5.0 years for TC (docetaxel and cyclophosphamide) of 86% vs. 80% for AC (doxorubicin and cyclophosphamide) (HR 0.67; 95% CI 0.5-0.94; P=0.015)(Figure 2). The purpose of this trial was to compare the clinical outcomes in patients treated with a standard adjuvant anthracycline regimen vs. a non-anthracycline regimen. Virtual simulations by the model correctly predicted that adjuvant TC (Docetaxel and Cyclophosphamide) would be superior to adjuvant AC (doxorubicin and cyclophosphamide) in early breast cancer (Predicted log odds ratio −0.059, P < 0.001; Table 3)

##### Study 8 (Japan & South Korea)

To further test the limitations of the Digital Twin’s performance, we aimed to challenge it further. We tested it across mixed populations who received different neoadjuvant chemotherapy regimens containing either an anthracycline, a taxane, or both, and then subsequently received heterogeneous adjuvant therapy. CREATE-X (Masuda et al.^54^ 2017) was a Phase 3 prospective, randomised study (n = 910) that randomised participants with residual disease following different neoadjuvant chemotherapy regimens for breast cancer (stage I-III) to either capecitabine or a no capecitabine. The study participants included both hormone-positive (ER+ve, HER2-ve) and triple-negative (ER-ve, HER2-ve) patients. For simulation purposes, we focused on the hormone-positive subpopulation only and assumed participants in the control arm would receive endocrine treatment but no capecitabine. For the CREATE-X study, the overall survival 95% confidence intervals and hazard ratios for the hormone-positive subgroup crossed 1.0 (n=601; HR 0.73 0.38-1.40; P = 0.41) suggesting adjuvant capecitabine was no better control. Virtual simulations by the Digital Twin model predicted that in people with hormone-positive breast cancer (HER2-ve; stages 1-3), treatment with adjuvant capecitabine would be inferior to standard of care such as adjuvant hormone treatment with tamoxifen (log odds ratio = 0.07). Although the predicted confidence intervals inferred inferiority, the prediction was within the confidence interval of the actual trial results.

### Predicting survival

An Overall Survival (OS) model was integrated into the Digital Twin clinical trial simulator using a Random Survival Forest (RSF)^58^, a statistical non-parametric ensemble learning method. The learning target is time-to-event and event (censored/deceased) data, and primary output is survival probability vs. time curves.

Clinical data from 10,913 patients was pre-processed to yield a dataset comprising 4029 patients, with ages ranging from 11 to 90 years, spanning 23 different cancer types. This dataset along with the RECIST response categories from the TRM stage was used as input for the OS model.

We analysed five different prediction accuracy metrics: 1) cumulative dynamic AUC ROC, 2) Uno’s concordance index^59^ (C-index), 3) time-dependent Brier score, 4) the Brier skill score, and, 5) explained variance. Table 4 shows the average outcome across 5-fold training and testing splits. A detailed explanation of the metrics is provided in the Online Methods. Both the dynamic AUC and the C-index scores of the Digital Twin were above 0.7 in a pan-cancer setting, a threshold set for a good predictive model.

**Table 4.** shows pan-cancer Digital Twin overall survival performance evaluation metrics. Overall, we observe relatively high accuracies evaluated across all types of cancer tissue types. The metrics shown are the average of 5-fold train and test splits.

| AUC ROC | C-index | Brier score | Brier skill | Explained variance |
| --- | --- | --- | --- | --- |
| 0.78 | 0.71 | 0.168 | 0.42 | 0.44 |

Additionally, we evaluated the performance of our Digital Twin OS prediction in relation to existing computational models found in the literature. The results are shown in Figure 3, with further details on these benchmarks available in Extended Data Table 1. When evaluating across all tissue types, the Digital Twin model exhibited commendable performance in comparison to the mean of all 29 other model-data methods and tissue types identified in the literature^60–77^. Regarding Glioma, our model demonstrated favourable performance compared to XGBoost-Surv by Dal Bo et al.^69^ (2023) and Deep Learning with Cox proportional hazard (CPH) by Jiang et al.^77^ (2021) on the Udine Hospital and TCGA datasets, respectively. In Breast Cancer and Glioblastoma our model performed comparably. Our model underperformed benchmarks for Ovarian Cancers and Head and Neck Cancers. This may be attributed to the absence of crucial data inputs, specifically, TNM cancer staging data.

**Figure 3:**
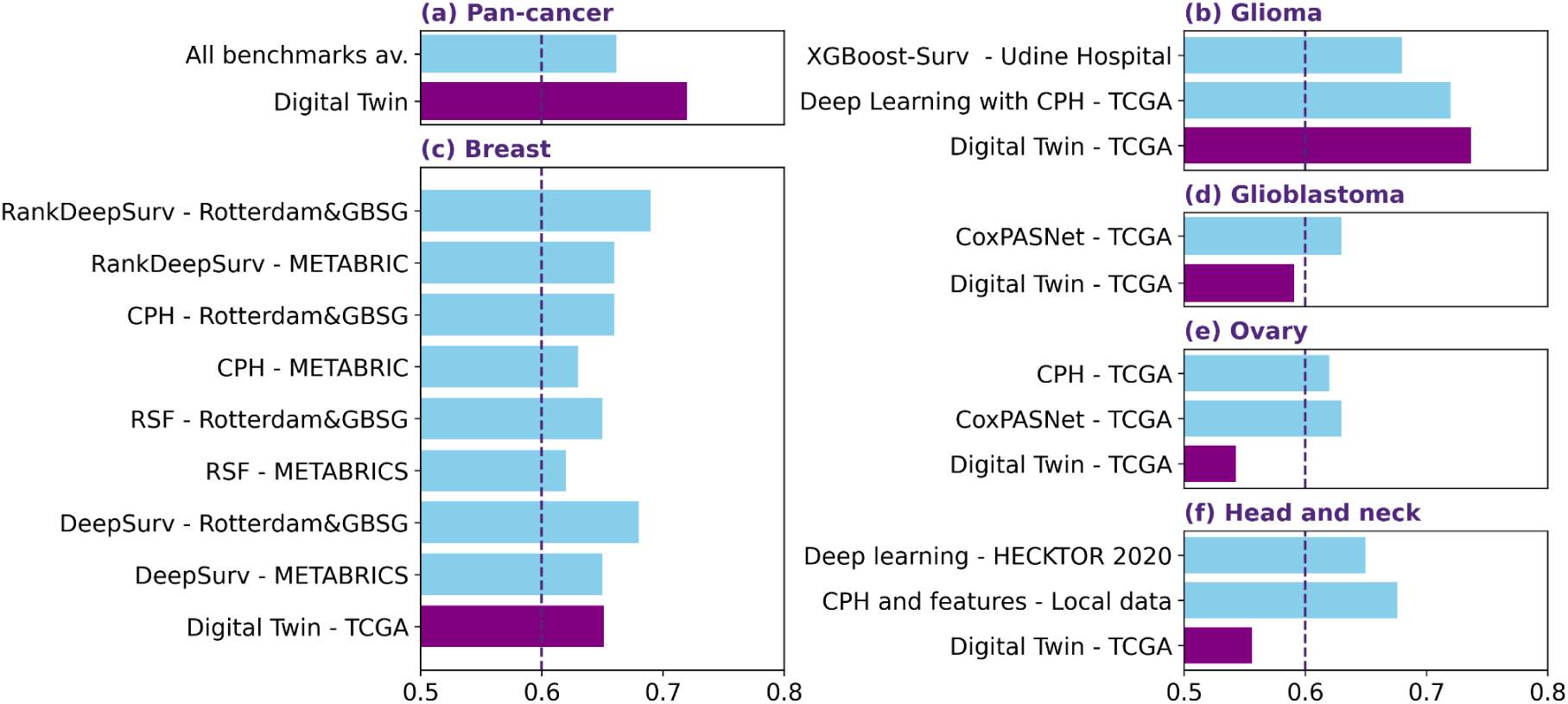
Benchmarking accuracy of overall survival method against existing methods. (a) A comparison of the C-index for 29 method-data benchmark approaches in modelling survival within the field of oncology, compared against our Digital Twin OS model for pan-cancer. (b-f) A similar comparison per specific cancer type. Details can be found in Extended Data Table 1.

However, benchmark studies typically do not address the prediction of survival curves for various drugs, including novel ones, and primarily focus on predicting survival curves for specific cancer types. In contrast, our model possesses the capability to address hypothetical scenarios, offering insights into questions such as the projected survival curve when a specific patient undergoes treatment with a novel drug.

### Cohort Enrichment

The FDA defines cohort enrichment as the “prospective use of any patient characteristic to select a study population in which detection of a drug effect (if one is in fact present) is more likely than it would be in an unselected population.” Enrichment strategies should accelerate drug development, increase the magnitude of drug responses and therefore accelerate the path to drug approval.

Here we evaluated the effectiveness of using the predicted response score to segment cohorts into responder and non-responder cohorts. For each cohort we chose two thresholds to split the cohort into positive, intermediate and negative groups and evaluated the log odds ratio of overall response rate to assess the potential for cohort enrichment.

The cohorts were split into 3 groups to assess the interpretability and quality of risk stratification of the response scores predicted by the model.This cohort molecular enrichment approach was tested in 19 different solid tumour types and 17 different cytotoxic drugs (Figure 4). The Digital Twin output data suggests that 16 solid tumour types, except ovarian cancer, would benefit from a molecular predictive enrichment strategy integrated into clinical trial designs. This data also confirms that machine learning approaches can successfully identify molecular patterns and enrich drug response for commonly used chemotherapy drugs such as docetaxel or cisplatin, which currently do not have robust predictive biomarkers and are used in unselected cancer populations.

**Figure 4.**
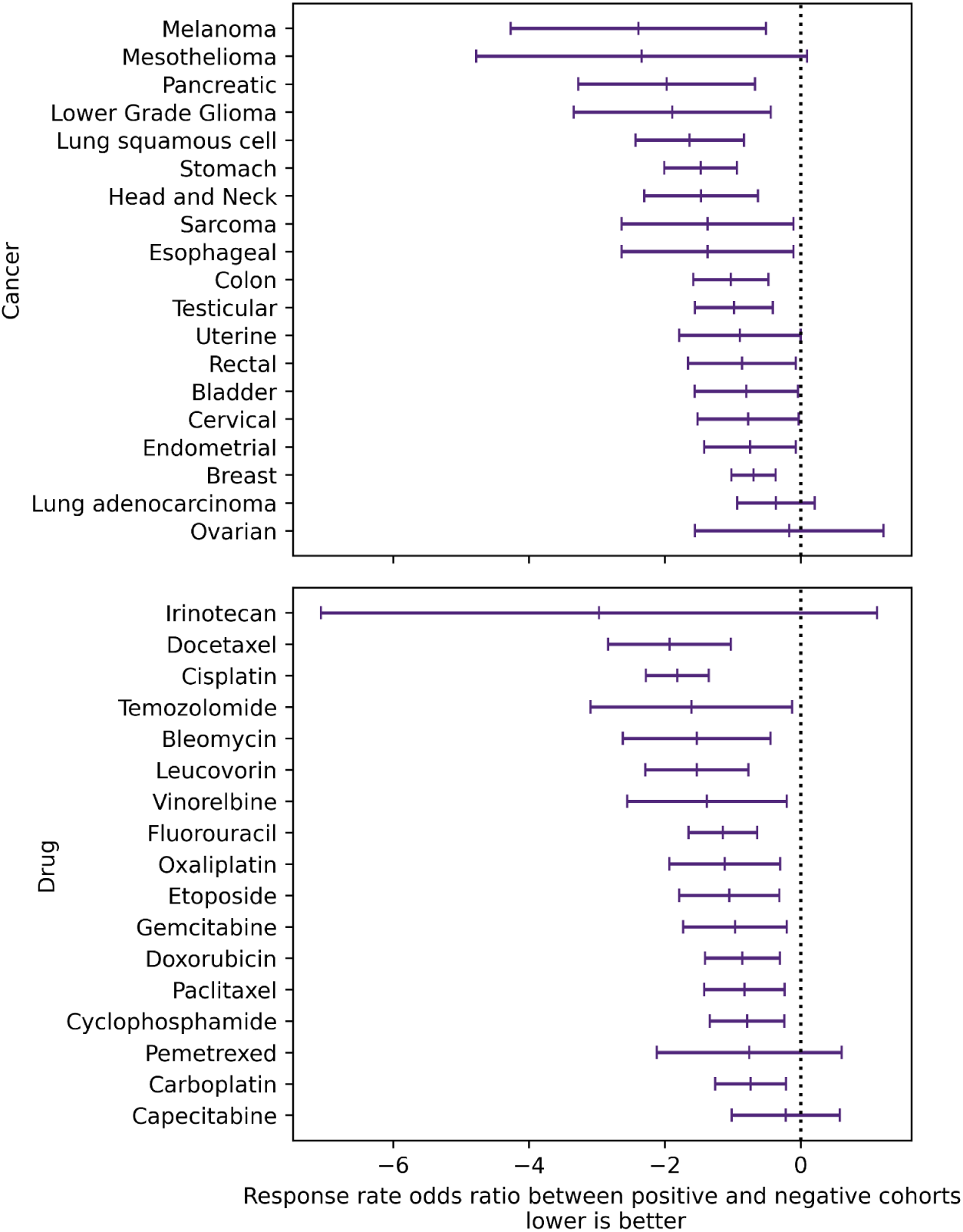
Assessing log odds increase in response rate through cohort enrichment. Cohort enrichment improves predicted therapeutic response rates showing that all cancers, perhaps with the exception of ovarian cancer would benefit from a molecular enrichment strategy to select responders. Similarly, with the exception of Irinotecan, pemetrexed and capecitabine the model did identify a significant benefit using the molecular data used to enrich for responders given the available patient numbers. The odds ratio in observed response rates between the biomarker positive and negative cohorts as segmented by treatment response predictions. The 95% confidence intervals are shown for each drug and cancer type.

## Discussion

Predicting therapeutic response has the potential to transform cancer care and oncology therapeutic development. We developed a Bayesian statistical approach that is similar to modelling gravitational lensing to predict the mass of dark matter by astrophysicists, where the complexity and interactivity of multiple data points is required. We show that this approach predicts with reasonable accuracy the response of therapeutics preclinically and clinically which we validated through comparison to clinical trials.

This approach can be applied at various points in therapeutic development. These include, but are not limited to:

1. Predicting response in preclinical models to inform decisions regarding which cancer and specific indication is more likely to be associated with response;
2. Combination strategies;
3. Selecting which therapeutics to advance through early clinical development;
4. Selecting which therapeutic to advance to late-stage development;
5. Improved patient selection for clinical development;
6. Construction of synthetic controls for clinical trials. In addition, other potential applications, not tested here, include prediction of toxicity.

One of the aspects of this approach is that the model can predict likely response for individual patients as well as cohorts. Predicting an individual’s response to a specific treatment ahead of time has the potential to substantially impact on routine cancer care. This would better inform clinical decision-making for an individual patient, avoiding likely ineffective therapies, and selecting the better option or a clinical trial. Moreover, increasing the accuracy of prediction for an individual would mean that they could potentially serve as their own control in a clinical trial. If the survival of an individual patient with standard of care could be predicted with a known level of accuracy, more meaningful information could be drawn from that individual’s response or lack of response to a novel therapy.

An important factor to consider is how accurate a prediction needs to be in order for it to be useful. With the high failure rate of oncology therapeutic development, an incremental increase in predictive accuracy for critical decisions would have potential significant impact on the probability of success.

Important current limitations that need to be addressed are the variability in predictive accuracy between different classes of therapeutics and different cancer types. Whilst this may be simply the amount and quality of data ingested, adjustments to the model may need to be made to reflect the mechanism of action of therapeutics, where known. Biological inferences from the model as it stands need to be developed further so as to better define candidate biomarkers that could be rapidly translated into the clinic.

## Data Availability

All data produced in the present study are available upon reasonable request to the authors

## Acknowledgements

This work was supported by Innovate UK [grant number 50074]. The data used in this study is in whole or part based upon data generated by the <u>TCGA Research Network</u>.

## Online Methods

Here, we describe a Digital Twin, an integrated machine-learning model for simulating clinical trials and estimating clinical trial endpoints for various treatment scenarios including novel drugs. It consists of three main models: the Drug Efficacy Model (DEM), the Treatment Response Model (TRM), and the Overall Survival (OS) model and employs multi-modal data input as illustrated previously in Figure 1. The DEM is trained to estimate the impact of treatment. It provides an estimation of a patient’s treatment response in the prospective cohort under the prospective treatment, entirely from preclinical data. With treatment response estimation, the overall survival model predicts the clinical trial endpoints.

### Drug Efficacy Model (DEM)

The DEM learns to estimate half-maximal inhibitory concentration (IC_50_) based on the pre-clinical information from cell-line studies. The model takes a set of features for the drug-patient pair: Whole Exome Sequencing (WXS), RNA sequencing (RNAseq), drug structures, and dose-response curves. The input data includes 545 compounds using SMILES (simplified molecular-input line-entry system) describing the structure of chemical species, 1343 genes in RNAseq, 1342 copy number variation (CNVs) from the WXS data and 130,000 Hill parameters for the dose–response curves. The drug structures were encoded as CDK^8–10^ molecular descriptors from their SMILES ID using cinfony^12^. The description of the input data for the Digital Twin DEM is shown in Extended Data Figure 1a.

Here, we deliberately opted for Random Forest^78,79^ (RF) because it allowed us to examine the leaf assignments that create the IC_50_ estimates. These leaf assignments are later used in the estimation of treatment response categories in the next stages of Digital Twin. Besides, RF tends to outperform other models in prognostic value^80^. DEM produces three types of outputs: 1) an estimate of the IC_50_ of a given drug in the given patient’s tissue, 2) drug combination synergy curves (the phenomenon where the combined effect of two or more drugs is greater than the sum of their individual effects), and 3) Perturbation Kernel^81^ leaf assignments. The latter is the main output, and input to the next stage of the Digital Twin, to the Treatment Response Model.

To evaluate the performance of the DEM, i.e. evaluate how good the predictions of a model on unseen validation data, we compare predicted against observed IC_50_ in Extended Data Figure 2. The root mean squared error (RMSE) of the log_10_ IC_50_ was 0.47 the coefficient of determination was 0.61.

### Perturbation Kernel

Here, a Perturbation Kernel is a method based on a Random Forest Kernel (RFK)^81^, that helps find patterns and relationships between data points in a more complex space where those patterns might be more apparent. It is constructed using the random partition sampling scheme, generating several RF decision trees, which are trained on a subset of features. In trained RF, each leaf in each tree is considered a partition. In learning algorithms, a kernel is a function that calculates the similarity or distance between pairs of data points. It is calculated based on counting the fraction of times these data points share a partition. Thus, the more these points share the same partition, the less distance between them and the more similar they are.

The Perturbation Kernel leaf assignments are output derived from the DEM and are utilised to transfer knowledge concerning shared molecular mechanisms across in vitro studies, combination screens, and clinical treatments. It outlines the similarity in molecular mechanisms between pairs of patients, for example, a patient treated with a taxane and another patient treated with an anthracycline. Subsequently, these are employed to train the Treatment Response Model for the prediction of treatment response categories.

### Treatment Response Model (TRM)

The TRM learns to accurately predict four RECIST treatment response categories^49^ from the output of DEM: 1) clinical progressive disease 2) stable disease, 3) partial response 4) complete response. The input to the model are the leaf assignments from the Perturbation Kernel, that is the output from the DEM. Random Forest in DEM allows the use of leaf assignments to form a kernel function between inputs. Assuming that similarity in IC_50_ estimates correlates with similarity in treatment response, it was possible to conduct kernel regression to estimate treatment response categories. To perform kernel regression we use the Gaussian Process^2^.

The performance of the TRM was evaluated based on its ability to classify the response categories in the overall pan-cancer setting, considering individual cancer types and distinct cancer treatments (see Extended Data Table 2). The following grouped RECIST categories were analysed: 1) Disease Control (combined complete response, partial response and stable disease) 2) Response (combined complete response and partial response) and 3) Complete Response. Overall, TRM demonstrated high accuracy, scoring 0.75, 0.74 and 0.69 in the respective grouped categories, along with Area Under the Curve (AUC) values of 0.63, 0.73, and 0.72. AUC reports the Receiver Operating Characteristic area under the curve for the non-thresholded prediction, and can be interpreted as the probability that a positive responder had a higher predicted probability of response than a negative responder. These were evaluated in a 5-fold cross-validation, and the reported value is the mean across the held-out cross-fold validation sets.

In addition, Extended Data Table 2 shows detailed model performance evaluation. Specifically, it displays accuracy, AUC, precision, recall and F1 score across a) grouped RECIST categories b) 9 cancer types and c) 12 cancer drugs. The Weighted Average provides scores weighted by the support for positive and negative responses. Average precision is the area under the precision-recall curve, measuring the average precision overall of all classification thresholds as a function of recall, with higher values being preferable.

Significant variations in performance are evident across treatments and tissue types, and much of this variability is likely attributed to the limited availability of response data for many cohorts.

### Statistics

The clinical trials confidence intervals are calculated according to Altman^82^ and Sheskin^83^. To calculate the predicted log(OR) (log odds ratio), the log(OR) was calculated for each individual patient in the dataset based on the treatment response model’s prediction, and the mean log(OR) and standard error were directly calculated. Because the log(OR) can be calculated for each individual patient the confidence intervals are much smaller than for an equivalent clinical study which is measuring the difference in response between two *populations*. When calculating the log odds Overall Response Rate (ORR) for a combination therapy the highest log odds for all treatments in the combination was taken.

### Overall Survival

We built an Overall Survival (OS) model as an integral component of the Digital Twin clinical trial simulator. A survival model is a statistical method to predict the time until an event, such as death. It deals with the censored data, indicating that the event of interest did not happen during the study period, and produces survival probability vs. time curves. We employed Random Survival Forest (RSF), a non-parametric ensemble learning method that can incorporate censored and time-to-event data^58^. The learning process involves the creation of multiple decision trees, and the model is selected based on the accuracy of predictions on unseen data.

The inputs for the OS are: 1) clinical records data, 2) cancer tissue type and 3) RECIST response categories from the TRM stage of the Digital Twin. Initially, 10,967 patients with survival data were pre-processed to end up with data from 4,029 patients with cancer tumour stages 0-4, lymph node stages 0-3, metastasis status (yes/no), and ages ranging from 11 to 90 years, across 23 cancer tissue types. Cancer types with small amounts of patients were removed from the analysis.

For patients with missing data, missing values were imputed with the mean values. The imputation was performed separately in the train set and validation set. Here, we group RECIST response categories into binary: 1) Disease (clinical progressive disease and stable disease, and 2) Response (partial response and complete response). The descriptive figure about the data is shown in Extended Data Figure 1.

To evaluate the performance of the model, we analysed five different prediction accuracy metrics across the solid tumours: 1) area under the receiver operating characteristic (AUC ROC) averaged for all times also known as cumulative dynamic AUC^84^ and 2) Uno’s concordance index (C-index) based on the inverse probability of censoring weights^59^. It is a goodness of fit measure for models that produce risk scores, commonly used in survival analysis. The intuition behind the C-index is – when comparing patients against each other if the patient with the higher risk score has a shorter time-to-event; 3) the integrated time-dependent Brier score^85^. It provides an overall calculation of the model performance at all available times. The smaller numerical values represent higher prediction accuracy (0 is the best achievable score with perfect accuracy and 1 is the worst score). 4) the Brier skill score that is the difference between the Brier score of the reference mode and the Brier score of the forecast model, divided by the Brier score of the reference model (1 is the best achievable score) and 5) explained variance, which is a proportion to which a model accounts for the variation of a given data set. Time-dependent metrics are integrated over time. We used scikit-survival^86^ for calculating the majority of metrics.

We conduct the performance evaluation using a 5-fold split (train/test splits), and all survival metrics reported are averaged across all cross-folds. In the pan-cancer setting, the model attained high accuracies as described by the following: AUC ROC = 0.78, C-index = 0.71, Brier score = 0.168, Brier skill = 0.42, and Explained Variance = 0.44. The performance of the OS per cancer was shown in Table 5 of the main body.

For the comparison with the benchmark studies (Figure 3 in the main body), we conducted a literature survey, selecting studies that focus on the survival analysis modelling within the oncology field. The complete list of studies is available in Extended Data Table 1. Given that the majority of studies utilise Harrell’s C-index^88^, we computed and compared it on a per-cancer basis.

### Feature Importance

We performed the feature importance analysis which emphasised the fact that the predicted output from the DEM modulates the Overall Survival, influencing clinical trial outcomes.

We used two methods: permutation-based importance and Cox proportional hazards model. The former is calculated by randomly permuting a feature and measuring the difference between the model prediction score after permutation and without. Cox estimates the impact of individual covariates on the hazard ratio (HR), allowing to quantify how changes in specific features affect the risk of an event occurring over time. The latter is a standard survival analysis model in oncology^87^.

Results are shown in Extended Data Fig 3. Results indicate that the “Disease (No response)”, which is inferred from the DEM plays a significant role in model performance. A stronger “disease” results in a positive log(HR) on the Cox model with the highest HR among all features. The permutation importance method shows that this feature has high importance, almost comparable to the patient’s age, which is considered one of the most important factors in cancer survival.

### Integration

Finally, we integrate DEM, TRM and OS models in order to be able to simulate clinical trials for both existing and novel cancer therapies. We call it a Digital Twin, a virtual model designed to accurately reflect clinical trials of cancer treatments with cytotoxic and small molecule therapies across various cancer types. The simulation output can then be any desired endpoint as predicted by the overall survival model.

The integration of distinct models involves utilising outputs from the preceding model as inputs for the subsequent model. DEM incorporates pre-clinical data, including gene expression and mutation profiles, drug response curves and drug compounds and produces a perturbation kernel. This perturbation kernel is subsequently employed in the TRM for predicting RECIST response categories. These predicted response categories are then incorporated into the OS model, along with clinical records data, to forecast overall patient survival over time.

---

**Extended Data Figure 1:**
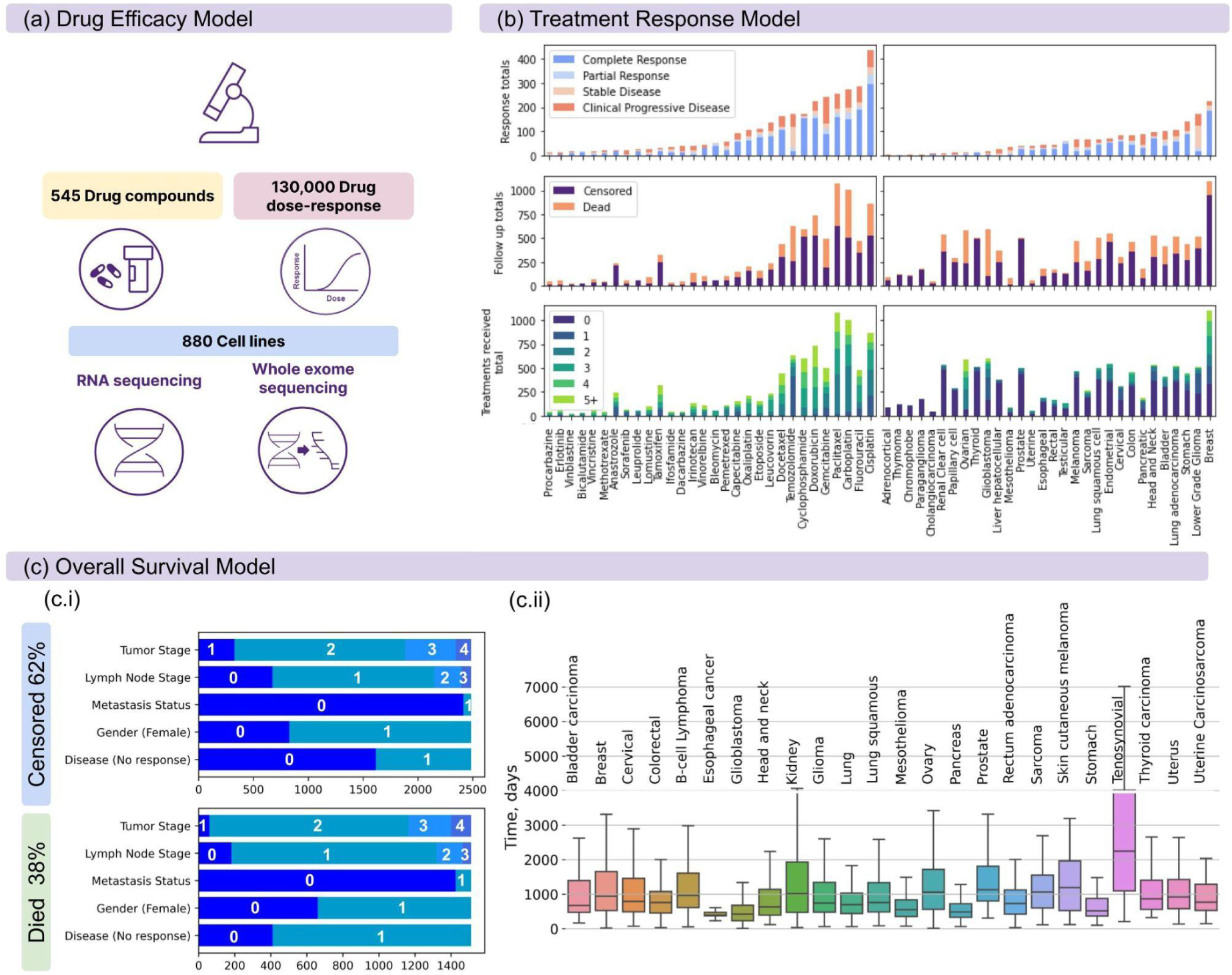
Digital Twin input data description. Description of the input data used to train the Drug Efficacy Model (a), Treatment Response Model (b) and Overall Survival Model (c). The distribution of clinical features across the event type (died/censored) (c1). The distribution of time-to-event data across cancer types (c2).

**Extended Data Figure 2.**
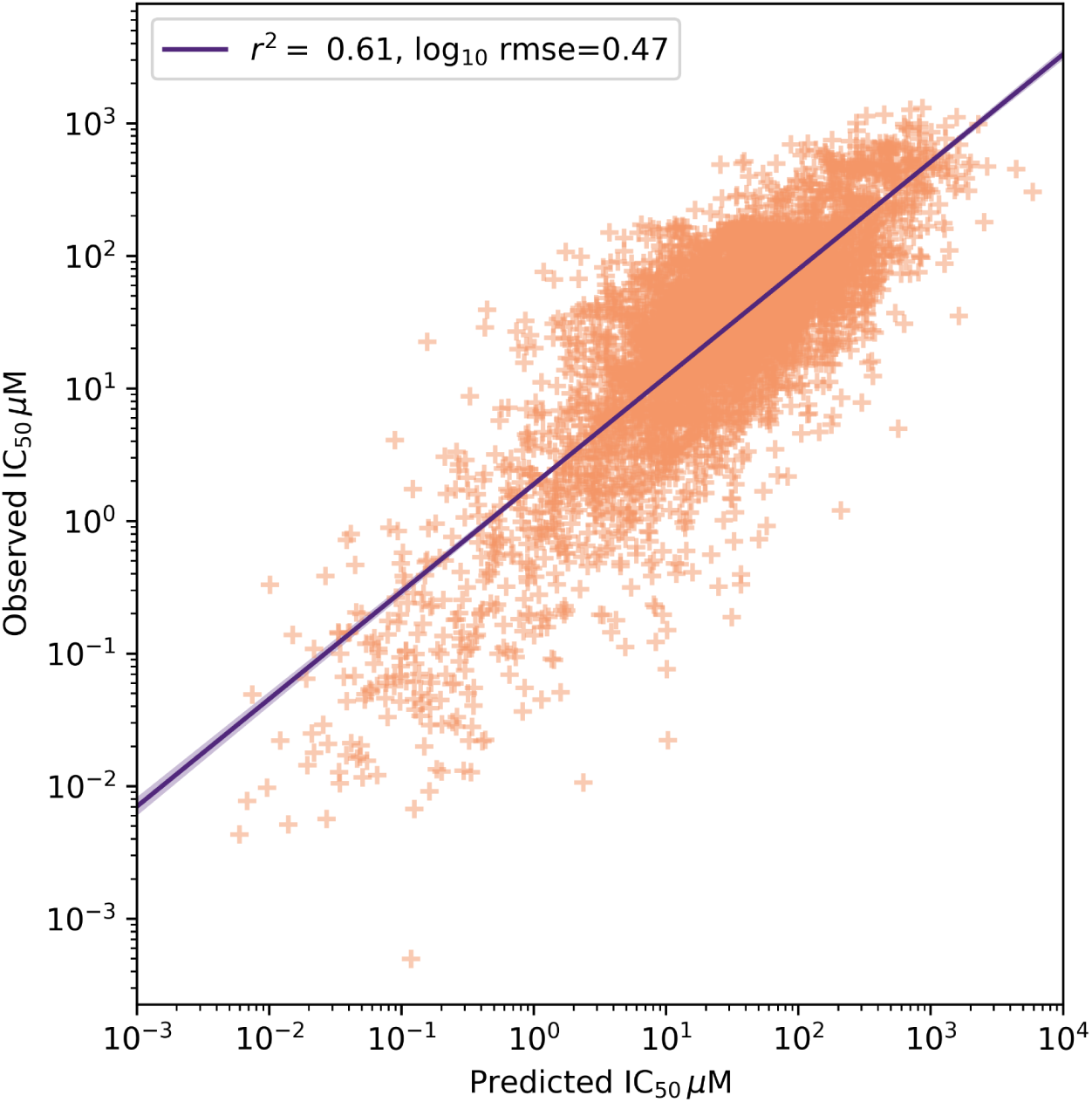
Drug Efficacy Model Performance. We compare observed vs predicted IC_50_ to evaluate the performance of the Drug Efficacy Model. The root mean squared error (RMSE) of the log10 IC50 was 0.47 the coefficient of determination was 0.61.

**Extended Data Figure 3:**
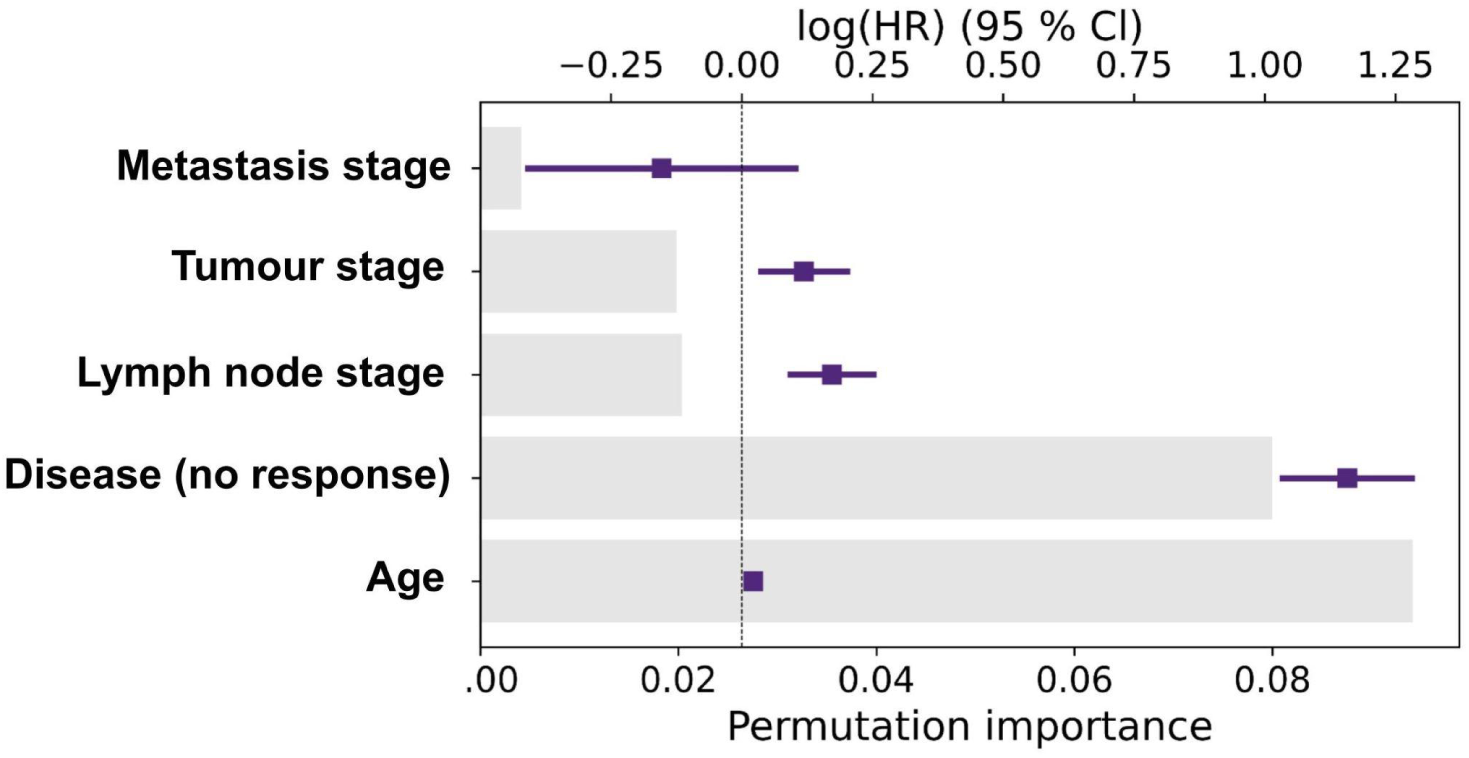
Feature importance. Feature importance: permutation-based importance (lower x-axis) and Cox hazard ratio-based importance (upper x-axis). Results indicate that the “disease (no response)” feature, which is inferred from the DEM plays a significant role in model performance. Results are based on a single split.

**Extended Data Table 1:** Overall survival comparisons. C-index metrics for benchmark computational models, with a focus on survival analysis in oncology. Time-dependent survival area under the curve (AUC) is infrequently reported in scientific publications. There are two concordance indices: defined by Harrell et al., (1984)^88^ and by Uno et al. (2011)^59^ which is based on the inverse probability of censoring weights. We utilise both of them, as the latter does not overestimate the index when there are a small number of events. However, the majority of studies use Harrell’s C-index.

| Method-Data | C-index | Notes | Reference |
| --- | --- | --- | --- |
| DeepSurv - METABRIC, Rotterdam&GBSG, SUPPORT | 0.65, 0.68, 0.62 | METABRIC - The Molecular Taxonomy of Breast Cancer International Consortium (METABRIC). GBSG - German Breast Cancer Study Group (GBSG). Rotterdam - is a breast cancer data set. SUPPORT - Study to Understand Prognoses Preferences Outcomes and Risks of Treatment (cancer is present as a feature only). Harrell C-index is used. Only breast cancer. | Katzman et al. (2018), Curtis et al., (2012), Royston and Altman (2013), Schumacher et al., (1994). Knaus et al., (1995). |
| RSF - METABRIC, Rotterdam&GBSG, SUPPORT | 0.62, 0.65, 0.62 |  |  |
| CPH - METABRIC, Rotterdam&GBSG, SUPPORT | 0.63, 0.66, 0.58 | CPH - cox proportional hazards model. |  |
| CPH for survival and Deep Learning for identification of tissue type - TCGA | 0.62 | RH - radiomic-histopathological combining modalities mode is the highest c-index. Focus on ovarian cancer (OV). Multimodal data. | Boehm et al. (2022) |
| AECOX - TCGA | 0.66 | AutoEncoder with Cox regression network. Train and evaluate 12 cancer types. Type of c-index not specified. | Huang et al., (2020) |
| CoxPASNet - TCGA | 0.63 | Only two cancer types are analysed - Glioblastoma and ovarian. Use TCGA genomic and clinical data. The comparison is performed against Cox-EN, Cox-nnet, and SurvivalNet. | Hao et al., (2018) |
| MesoNet - MESOBANK and TCGA | 0.64-0.65 | Predict survival of mesothelioma patients from digitised images. | Courtiol et al., (2019) |
| RankDeepSurv - METABRIC, Rotterdam&GBSG, SUPPORT | 0.66, 0.69, 0.62 | A deep feedforward neural network with new loss function, defined as the summation of an extended mean squared error loss and a pairwise ranking loss, incorporating survival data-based ranking information. Only breast cancer. | Jing et al., (2019) |
| LSTM - SCap | 0.81 | Prostate cancer only. Harrell's C-index is used. AUC is provided 0.83. | Koo et al., (2020) |
| CPH - MSKCCPAN | 0.62 | Uno's c-index used. MSKCCPAN - Memorial Sloan Kettering Cancer Center Pancreatic Adenocarcinoma Nomogram. This is a prediction tool to help physicians make treatment decisions. Pancreatic cancer only. | Marcinak et al., (2023) |
| Nomogram | 0.71 | A nomogram for predicting the occurrence of postoperative colorectal lesions in colorectal cancer. | Kawai et al., (2015) |
| SPSS software and STATA/SE 10 - Local data | 0.72 | The paper is analysing an optimal TNM staging edition and criteria. The c-index (Harrell's) is taken as the average of all stages present TNM5 - TNM7. Colorectal cancer. | Ueno et al., (2012) |
| Deep learning - HECKTOR 2020 | 0.672, 0.626 | C-index is for Hand-Crafted (HC) radiomics features and fully automatic pipeline respectively. Head and Neck cancer. | Fontaine et al., (2021) |
| Feature extraction with CPH - Local German dataset | 0.78, 0.61, 0.64 | CT, and two FDG-PET features models respectively. | Starke et al., (2023) |
| Deep Learning with CPH -TCGA | 0.784, 0.654 | Lower-grade gliomas. The 0.654 c-index is the WHO grade 3 gliomas from TCGA dataset, which were not used for training. | Jiang et al., (2021) |
| XGBoost-Surv - Udine Hospital data | 0.68 | Glioma grade 4. Clinical data, radiological data, or panel-based sequencing data such as presence of somatic mutations and amplification. | Dal Bo et al., (2023) |

**Extended Data Table 2:** Treatment Response Model Performance. A summary of the performance evaluation metrics of the treatment response model (TRM). The metrics were evaluated in a cross-fold validation with 5 splits. The reported value is the mean across the held-out cross-fold validation sets. The output of the TRM is a probability of response, so for the binary accuracy metrics, the prediction was thresholded such that the predicted response rates matched that of the training set.

|  |  | Metric |  |  |  |  |  |  |  | Positive |  |  | Weighted Avg. |  |  |
| --- | --- | --- | --- | --- | --- | --- | --- | --- | --- | --- | --- | --- | --- | --- | --- |
|  |  | Acc. | AUC | Avg.<br>precision | response<br>rate | PPV | NPV | Sensitivity | Specificity | Precision | Recall | f1-score | Precision | Recall | f1-score |
| Overall | Disease control | 0.747 | 0.625 | 0.883 | 0.830 | 0.849 | 0.261 | 0.846 | 0.257 | 0.849 | 0.846 | 0.847 | 0.750 | 0.747 | 0.748 |
|  | Response | 0.735 | 0.733 | 0.844 | 0.710 | 0.815 | 0.542 | 0.811 | 0.550 | 0.815 | 0.811 | 0.813 | 0.737 | 0.735 | 0.735 |
|  | Complete Response | 0.692 | 0.723 | 0.777 | 0.634 | 0.759 | 0.579 | 0.756 | 0.585 | 0.759 | 0.756 | 0.756 | 0.695 | 0.692 | 0.692 |
| Cancer | Lung squamous cell | 0.774 | 0.651 | 0.773 | 0.607 | 0.732 | 1.000 | 1.000 | 0.333 | 0.732 | 1.000 | 0.845 | 0.795 | 0.774 | 0.831 |
|  | Stomach | 0.669 | 0.542 | 0.814 | 0.736 | 0.750 | 0.333 | 0.824 | 0.238 | 0.750 | 0.824 | 0.785 | 0.640 | 0.669 | 0.696 |
|  | Head and Neck | 0.643 | 0.424 | 0.750 | 0.786 | 0.750 | 0.000 | 0.818 | 0.000 | 0.750 | 0.818 | 0.783 | 0.589 | 0.643 | 0.783 |
|  | Colon | 0.615 | 0.410 | 0.601 | 0.633 | 0.638 | 0.250 | 0.939 | 0.083 | 0.638 | 0.939 | 0.754 | 0.537 | 0.615 | 0.710 |
|  | Lung adenocarcinoma | 0.605 | 0.536 | 0.672 | 0.605 | 0.617 | 0.500 | 0.917 | 0.133 | 0.617 | 0.917 | 0.736 | 0.575 | 0.605 | 0.702 |
|  | Pancreatic | 0.541 | 0.527 | 0.563 | 0.469 | 0.500 | 0.564 | 0.373 | 0.700 | 0.500 | 0.373 | 0.418 | 0.544 | 0.541 | 0.526 |
|  | Bladder | 0.437 | 0.400 | 0.610 | 0.563 | 0.500 | 0.000 | 0.775 | 0.000 | 0.500 | 0.775 | 0.608 | 0.282 | 0.437 | 0.608 |
| Drug | Docetaxel | 0.787 | 0.565 | 0.839 | 0.860 | 0.871 | 0.500 | 0.853 | 0.500 | 0.871 | 0.853 | 0.860 | 0.792 | 0.787 | 0.785 |
|  | Cisplatin | 0.781 | 0.667 | 0.874 | 0.799 | 0.836 | 0.370 | 0.901 | 0.297 | 0.836 | 0.901 | 0.867 | 0.749 | 0.781 | 0.791 |
|  | Platinum therapies | 0.764 | 0.649 | 0.873 | 0.804 | 0.837 | 0.347 | 0.881 | 0.300 | 0.837 | 0.881 | 0.857 | 0.741 | 0.764 | 0.750 |
|  | Fluorouracil | 0.757 | 0.617 | 0.843 | 0.825 | 0.844 | 0.396 | 0.852 | 0.381 | 0.844 | 0.852 | 0.848 | 0.756 | 0.757 | 0.756 |
|  | Anthracyclines | 0.753 | 0.647 | 0.846 | 0.894 | 0.801 | 0.000 | 0.922 | 0.000 | 0.801 | 0.922 | 0.857 | 0.658 | 0.753 | 0.857 |
|  | Doxorubicin | 0.753 | 0.647 | 0.846 | 0.894 | 0.801 | 0.000 | 0.922 | 0.000 | 0.801 | 0.922 | 0.857 | 0.658 | 0.753 | 0.857 |
|  | All therapies | 0.732 | 0.695 | 0.852 | 0.759 | 0.825 | 0.446 | 0.823 | 0.454 | 0.825 | 0.823 | 0.823 | 0.734 | 0.732 | 0.732 |
|  | Taxanes | 0.712 | 0.608 | 0.844 | 0.776 | 0.805 | 0.374 | 0.833 | 0.314 | 0.805 | 0.833 | 0.816 | 0.712 | 0.712 | 0.706 |
|  | Paclitaxel | 0.667 | 0.542 | 0.774 | 0.727 | 0.746 | 0.393 | 0.825 | 0.274 | 0.746 | 0.825 | 0.779 | 0.660 | 0.667 | 0.649 |
|  | Carboplatin | 0.657 | 0.555 | 0.828 | 0.774 | 0.767 | 0.191 | 0.788 | 0.190 | 0.767 | 0.788 | 0.775 | 0.641 | 0.657 | 0.678 |
|  | Oxaliplatin | 0.650 | 0.617 | 0.776 | 0.806 | 0.700 | 0.500 | 0.783 | 0.433 | 0.700 | 0.783 | 0.736 | 0.635 | 0.650 | 0.636 |
|  | Gemcitabine | 0.512 | 0.566 | 0.705 | 0.624 | 0.664 | 0.411 | 0.471 | 0.604 | 0.664 | 0.471 | 0.539 | 0.579 | 0.512 | 0.516 |

## Notes

### Competing Interest Statement

The authors have declared no competing interest.

### Author Declarations

The human data used in this study is in whole or part based upon public data generated by the TCGA Research Network.

